# Time Metrics of Acute Stroke Pathways for Mechanical Thrombectomy in Indonesia: A Descriptive Study on Two Tertiary Major Hospitals

**DOI:** 10.64898/2026.07.14.26358067

**Authors:** Affan Priyambodo Permana, Sudarto Ronoatmodjo, Kevin Gunawan, Setyo Widi Nugroho, Mohammad Kurniawan, Al Rasyid, Radi Muharris Mulyana, Syahrul, Reza Aditya Arpandy, Yoga Arif Syah Hidayat, Khairunnisa Farina Ilato, Bryan Gervais de Liyis, Nur Afianti Hasanah, Asri Chasanah Adisasmita

**Affiliations:** Department of Neurosurgery, Faculty of Medicine, Universitas Indonesia, Dr. Cipto Mangunkusumo National General Hospital (RSCM), Jakarta, Indonesia; Faculty of Public Health, Universitas Indonesia, Depok, Indonesia; Department of Neurology, Faculty of Medicine, Universitas Indonesia, Dr. Cipto Mangunkusumo National General Hospital (RSCM), Jakarta, Indonesia; Emergency Unit Dr. Cipto Mangunkusumo National General Hospital (RSCM), Jakarta, Indonesia; Department of Neurology, National Brain Center Hospital Prof. Dr. dr. Mahar Mardjono (RSPON), Jakarta, Indonesia; Department of Neurosurgery, National Brain Center Hospital Prof. Dr. dr. Mahar Mardjono (RSPON), Jakarta, Indonesia

**Keywords:** mechanical thrombectomy, acute ischemic stroke, large-vessel occlusion, workflow metrics, stroke systems of care

## Abstract

**Background:** Mechanical thrombectomy (MT) is a time-sensitive reperfusion treatment for acute ischemic stroke caused by large-vessel occlusion. Workflow time metrics for MT remain poorly characterized in Indonesia, where stroke burden is substantial. This study describes pre-hospital and in-hospital time-interval metrics for MT across two major tertiary hospitals in Jakarta and evaluates institutional trends over a nine-year period.

**Methods:** We conducted a retrospective descriptive study of consecutive patients undergoing MT at dr. Cipto Mangunkusumo National General Hospital (RSCM) and Prof. Dr. dr. Mahar Mardjono National Brain Center Hospital (RSPON) from 2017 to 2025. Pre-hospital and in-hospital time-interval metrics were reported as median (interquartile range [IQR]) and stratified by institution.

**Results:** Among 330 registered patients, 71 were excluded due to incomplete data, leaving 259 in the final cohort (RSCM n=38; RSPON n=221). The pooled cohort had a mean age of 58.12 ± 11.09 years; 63.71% were male. Hypertension was the most prevalent vascular risk factor (53.67%). Median door-to-CT time was 9 minutes (IQR 18), door-to-decision 101 minutes (IQR 100), and door-to-groin puncture 272 minutes (IQR 152). Total ischemic time (onset-to-groin puncture) was 468 minutes (IQR 294). MT volume increased substantially over the study period, particularly after 2022 at RSPON, which also demonstrated progressive improvement in in-hospital workflow times. RSCM showed increasing delays in later years, consistent with institutional congestion at a general multispecialty center.

**Conclusions:** Early brain imaging was achievable at both centers; however, post-imaging delays particularly in CT-to-groin intervals, represent the dominant in-hospital bottleneck. Future quality-improvement efforts should prioritize decision-making, team mobilization, and pre-hospital coordination to reduce total ischemic time and improve access to reperfusion therapy.

## Introduction

Acute ischemic stroke caused by large-vessel occlusion is a time-critical neurological emergency. Mechanical thrombectomy (MT), with or without intravenous thrombolysis, is now a cornerstone of reperfusion therapy for eligible patients and is supported by contemporary international guidelines [1]. Crucially, the benefit of MT is strongly time dependent. Delays are associated with worse outcomes [2,3]. Modern stroke systems focus not only on patient selection but also on optimizing workflow intervals that are directly modifiable by hospital processes. Key in-hospital metrics commonly used to monitor the pathway include door-to-imaging, door-to-decision, and door-to-groin puncture, alongside thrombolysis-specific metrics such as door-to-needle time [4]. The need to optimize these time metrics is particularly relevant in Indonesia, where stroke remains a major public health problem and has been reported as a leading cause of disability burden [5]. Indonesia has introduced code stroke systems to accelerate hyperacute stroke care. For example, a structured code stroke system has been running in dr. Cipto Mangunkusumo National General Hospital (RSCM) prior to 2014. Several studies have described selected aspects of code-stroke performance, such as door-to-CT time [6]. However, evaluations specifically focusing on MT workflow response times remain limited [7,8]. Therefore, this multicenter descriptive study aims to characterize the hyperacute stroke workflow and response time for patients undergoing MT in tertiary Indonesian hospitals. By providing a structured baseline picture of MT workflow metrics at two tertiary centers in Jakarta, this study aims to support benchmarking, identify workflow bottlenecks, and inform targeted quality-improvement initiatives to reduce modifiable delays and improve timely access to reperfusion therapy [9].

## Methods

### Study population

We conducted a two-center retrospective descriptive study at two tertiary hospitals in Jakarta, Indonesia. The participating hospitals were RSCM, a multispecialty academic referral hospital, and RSPON, a national neuro-focused referral hospital. Eligible participants were consecutive patients with acute ischemic stroke who underwent endovascular mechanical thrombectomy during 2017 to 2025. Patients who received intravenous alteplase prior to MT were included. Patients were excluded if medical records lacked key data required to compute the main workflow time-interval metrics or if other essential variables were incomplete such that the case could not be reliably characterized. Only cases meeting completeness criteria were included in the final analysis.

### Data collection

Data were extracted from hospital medical records and radiology reports from 2017 through 2025 at both centers and entered into a standardized data-collection form. Hospital records were accessed for research purposes between February 10^th^, 2023 to January 22^th^, 2024 and March 6^th^, 2025 to June 20^th^, 2026. The authors had access to information that could identify individual participants during data collection. However, all data were de-identified prior to analysis. Variables collected were demographic and baseline characteristics such as age, sex, vascular risk factors, pre-stroke functional status, stroke severity before thrombectomy, and admission blood glucose, stroke onset and presentation features such as symptom onset time, last known well time, and presenting neurological symptoms, acute reperfusion treatments data such as intravenous thrombolytic agent administrations, imaging and angiographic variables such as ASPECT score and occlusion locations, workflow time-interval metrics related to patient arrival, onset of stroke, imaging, thrombolysis, decision making by physicians, and time of thrombectomy. Cases with insufficient timestamp documentation preventing computation of the key workflow intervals were excluded as per protocol.

### Statistical analysis

Analyses were primarily descriptive. Continuous variables were summarized as mean ± SD if normally distributed or median (min-max) if skewed. Categorical variables were summarized as counts and percentages. Workflow time-intervals typically exhibit right-skewed distribution and very sensitive to outliers. Therefore, they were summarized using median (IQR), consistent with common reporting practices in large stroke registries. It is additionally presented overall and stratified by hospitals to illustrate inter-center variation [1].

## Results

### Baseline Cohort and Clinical Characteristics

Among 330 patients who underwent MT for acute ischemic stroke at RSCM and RSPON during the study period, 71 were excluded because essential data were incomplete, leaving 259 patients for the final analysis (**Figure 1**). The pooled cohort had a mean age of 58.12 ± 11.09 years, with a male predominance of 63.71% (n=165). Baseline clinical characteristics were consistent across the study population, with a median National Institutes of Health Stroke Scale (NIHSS) score of 14 (IQR 4). The most prevalent vascular risk factor was hypertension (53.67%), followed by smoking history (25.48%) and diabetes mellitus (20.85%). Pre-stroke functional independence (mRS 0) was present in 84.56% of the total population (**Table 1**).

**Fig. 1.**
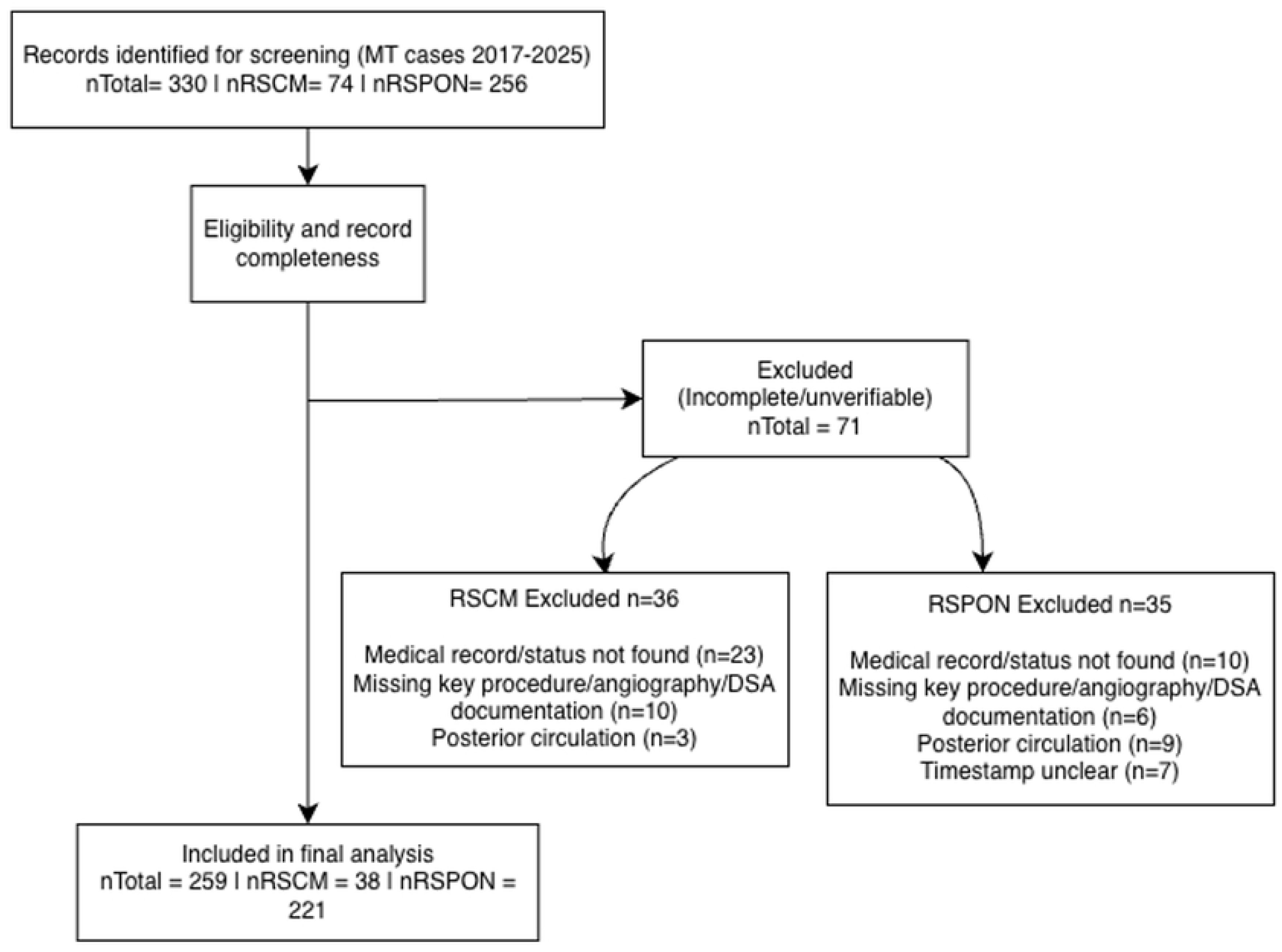
Study flow diagram of case identification, exclusions, and final study cohort inclusion across two tertiary hospitals in Jakarta (RSCM and RSPON), 2017-2025.

**Table 1.**
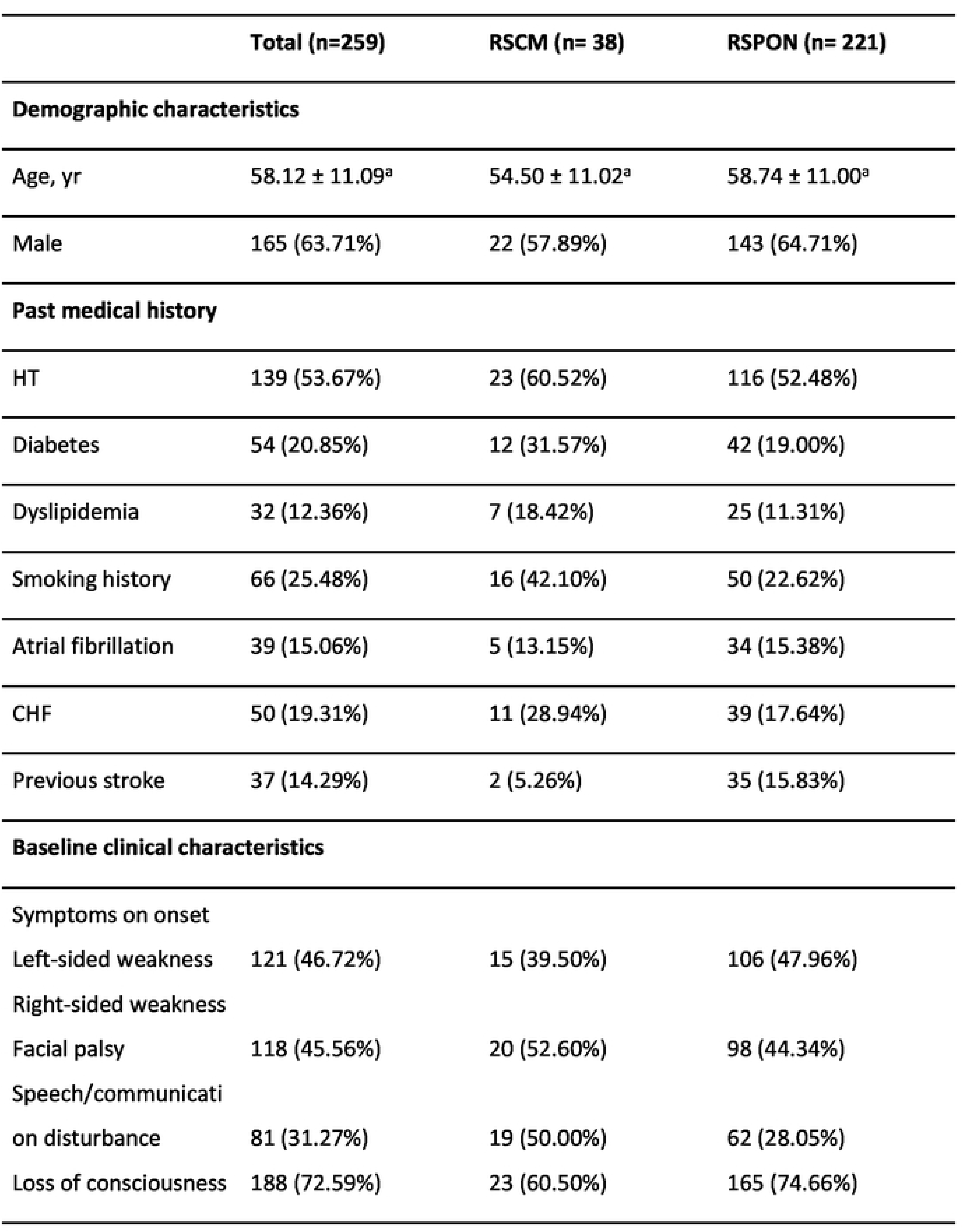
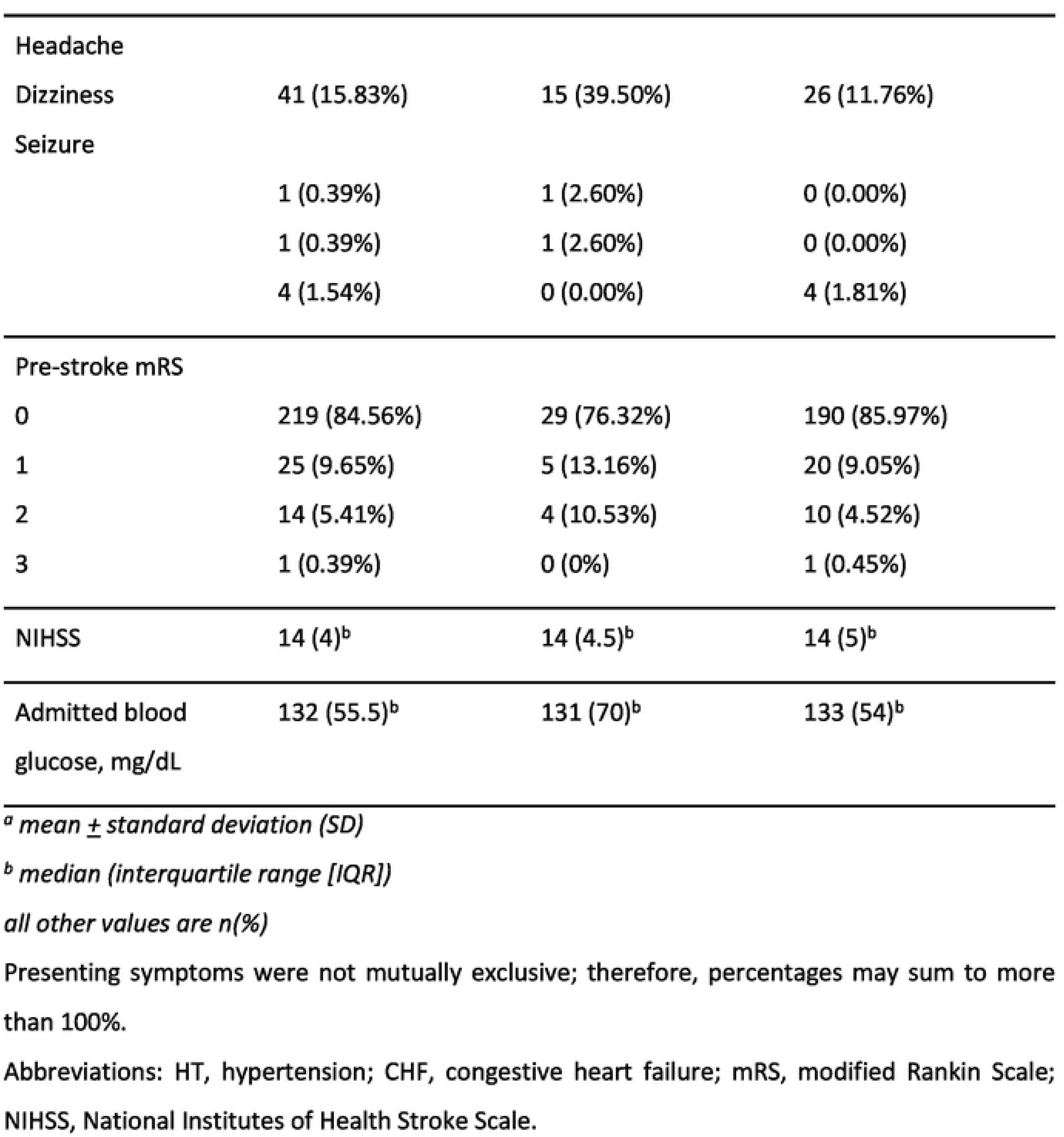
Demographic and baseline clinical characteristics of the total and stratified cohorts.

|  | Total (n=259) | RSCM (n= 38) | RSPON (n= 221) |
| --- | --- | --- | --- |
| **Demographic characteristics** |  |  |  |
| Age, yr | 58.12 ± 11.09<sup>a</sup> | 54.50 ± 11.02<sup>a</sup> | 58.74 ± 11.00<sup>a</sup> |
| Male | 165 (63.71%) | 22 (57.89%) | 143 (64.71%) |
| **Past medical history** |  |  |  |
| HT | 139 (53.67%) | 23 (60.52%) | 116 (52.48%) |
| Diabetes | 54 (20.85%) | 12 (31.57%) | 42 (19.00%) |
| Dyslipidemia | 32 (12.36%) | 7 (18.42%) | 25 (11.31%) |
| Smoking history | 66 (25.48%) | 16 (42.10%) | 50 (22.62%) |
| Atrial fibrillation | 39 (15.06%) | 5 (13.15%) | 34 (15.38%) |
| CHF | 50 (19.31%) | 11 (28.94%) | 39 (17.64%) |
| Previous stroke | 37 (14.29%) | 2 (5.26%) | 35 (15.83%) |
| **Baseline clinical characteristics** |  |  |  |
| Symptoms on onset |  |  |  |
| Left-sided weakness | 121 (46.72%) | 15 (39.50%) | 106 (47.96%) |
| Right-sided weakness |  |  |  |
| Facial palsy | 118 (45.56%) | 20 (52.60%) | 98 (44.34%) |
| Speech/communicati |  |  |  |
| on disturbance | 81 (31.27%) | 19 (50.00%) | 62 (28.05%) |
| Loss of consciousness | 188 (72.59%) | 23 (60.50%) | 165 (74.66%) |
| Headache |  |  |  |
| Dizziness | 41 (15.83%) | 15 (39.50%) | 26 (11.76%) |
| Seizure |  |  |  |
|  | 1 (0.39%) | 1 (2.60%) | 0 (0.00%) |
|  | 1 (0.39%) | 1 (2.60%) | 0 (0.00%) |
|  | 4 (1.54%) | 0 (0.00%) | 4 (1.81%) |
| Pre-stroke mRS |  |  |  |
| 0 | 219 (84.56%) | 29 (76.32%) | 190 (85.97%) |
| 1 | 25 (9.65%) | 5 (13.16%) | 20 (9.05%) |
| 2 | 14 (5.41%) | 4 (10.53%) | 10 (4.52%) |
| 3 | 1 (0.39%) | 0 (0%) | 1 (0.45%) |
| NIHSS | 14 (4)<sup>b</sup> | 14 (4.5)<sup>b</sup> | 14 (5)<sup>b</sup> |
| Admitted blood<br>glucose, mg/dL | 132 (55.5)<sup>b</sup> | 131 (70)<sup>b</sup> | 133 (54)<sup>b</sup> |

<sup>a</sup> mean ± standard deviation (SD)
<sup>b</sup> median (interquartile range [IQR])
all other values are n(%)
Presenting symptoms were not mutually exclusive; therefore, percentages may sum to more than 100%.
Abbreviations: HT, hypertension; CHF, congestive heart failure; mRS, modified Rankin Scale; NIHSS, National Institutes of Health Stroke Scale.

Regarding occlusion site, the middle cerebral artery (MCA) was the most commonly involved vessel (69.12%), followed by the Internal Carotid Artery (ICA) (27.80%). Baseline imaging severity was generally favorable. Nearly half of patients received IV thrombolysis prior to MT (45.60%) (**Table 2**).

**Table 2.** Imaging and treatment characteristics.

|  | Total (n=259) | RSCM (n= 38) | RSPON (n= 221) |
| --- | --- | --- | --- |
| **Imaging characteristics** |  |  |  |
| ASPECTS | 9 (3)<sup>b</sup> | 8 (3)<sup>b</sup> | 9 (3)<sup>b</sup> |
| Occlusion location |  |  |  |
| MCA | 179 (69.12%) | 30 (78.95%) | 149 (67.42%) |
| ICA | 72 (27.80%) | 6 (15.79%) | 66 (29.86%) |
| ACA | 7 (2.70%) | 2 (5.26%) | 5 (2.26%) |
| PCA | 1 (0.39%) | 1 (2.63%) | 0 (0.00%) |
| BA | 2 (0.77%) | 1 (2.63%) | 1 (0.45%) |
| CCA | 1 (0.39%) | 1 (2.63%) | 0 (0.00%) |
| VA | 2 (0.77%) | 0 (0.00%) | 2 (0.90%) |
| **Thrombolytic therapy prior to MT** |  |  |  |
| Yes | 118 (45.60%) | 15 (39.47%) | 103 (46.60%) |
| Dosage |  |  |  |
| 0.6 mg/kgBW | 60 (50.85%) | 14 (93.33%) | 46 (44.66%) |
| 0.9 mg/kgBW | 58 (49.15%) | 1 (6.67%) | 57 (55.34%) |

<sup>a</sup> mean ± standard deviation (SD)
<sup>b</sup> median (interquartile range [IQR])
all other values are n(%)
Occlusion-site categories were not mutually exclusive because some patients had tandem or multiterritory occlusions.
Abbreviations: ASPECTS, Alberta Stroke Program Early CT Score; MCA, middle cerebral artery; ICA, internal carotid artery; ACA, anterior cerebral artery; PCA, posterior cerebral artery; BA, basilar artery; CCA, common carotid artery; VA, vertebral artery; MT, mechanical thrombectomy.

### Trends in Mechanical Thrombectomy Volume

Over the nine-year study period, there was a marked increase in the total volume of MT procedures performed, but the trajectory differed substantially by institution. RSCM maintained a consistently low annual MT volume throughout the study period, whereas RSPON demonstrated a sharp expansion after 2022. Between 2017 and 2022, case volumes remained relatively stable and low, typically under 30 cases per year. This period of relative stagnation included the acute phases of the COVID-19 pandemic, which likely constrained hyperacute stroke services due to institutional resource redirection and pandemic-related healthcare disruptions, after which a marked post-pandemic surge was driven primarily by RSPON.

Following the end of the COVID-19 pandemic in 2022, a sharp upward trajectory in total MT volume was observed starting in 2023, peaking at over 100 cases in 2025, majorly in RSPON as the more neuro-specialized hospital (**Figure 2**). This post-pandemic surge reflects a combination of restored hospital capacity, increased public confidence in seeking emergency care, and the formal expansion of stroke pathways. This growth was primarily driven by RSPON, which experienced exponential volume increases in the final three years, while RSCM maintained a steady, lower case volume despite the overall national trend.

**Fig. 2.**
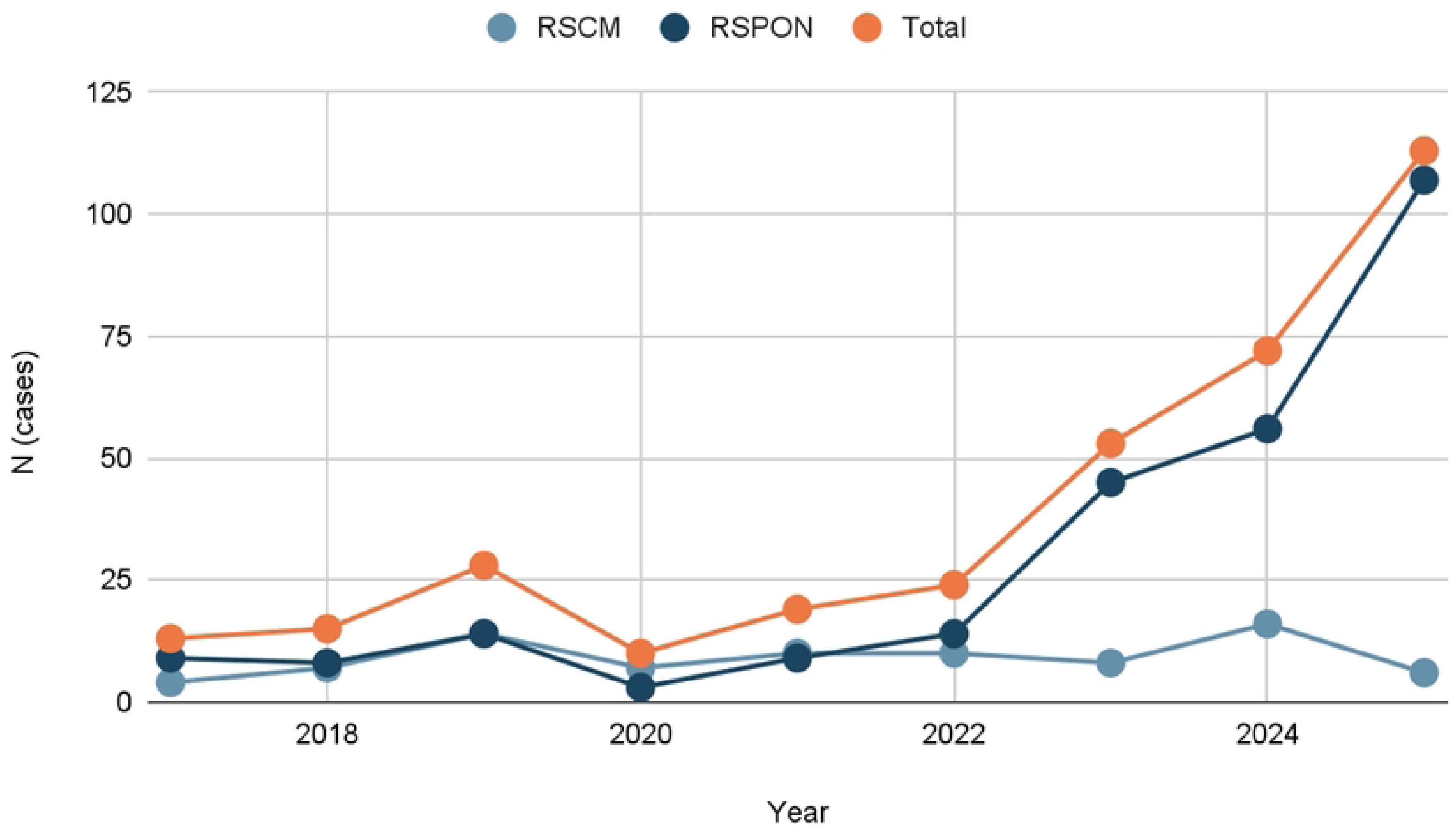
Annual trends in mechanical thrombectomy procedure volume.

### Aggregate Workflow and Time Metrics

The pooled median for in-hospital time metrics revealed a median door-to-CT time of 9 minutes (IQR 18) and a door-to-needle time of 55 minutes (IQR 74.5). The median time from hospital arrival to groin puncture (door-to-groin) was 272 minutes (IQR 152). Total ischemic time, measured from symptom onset to groin puncture, showed a pooled median of 468 minutes (IQR 294). While pre-hospital intervals (onset-to-door) were broadly comparable across the total cohort (median 170 minutes), significant variation was observed in the in-hospital workflow stages when stratified by institution (**Table 3**).

**Table 3.**
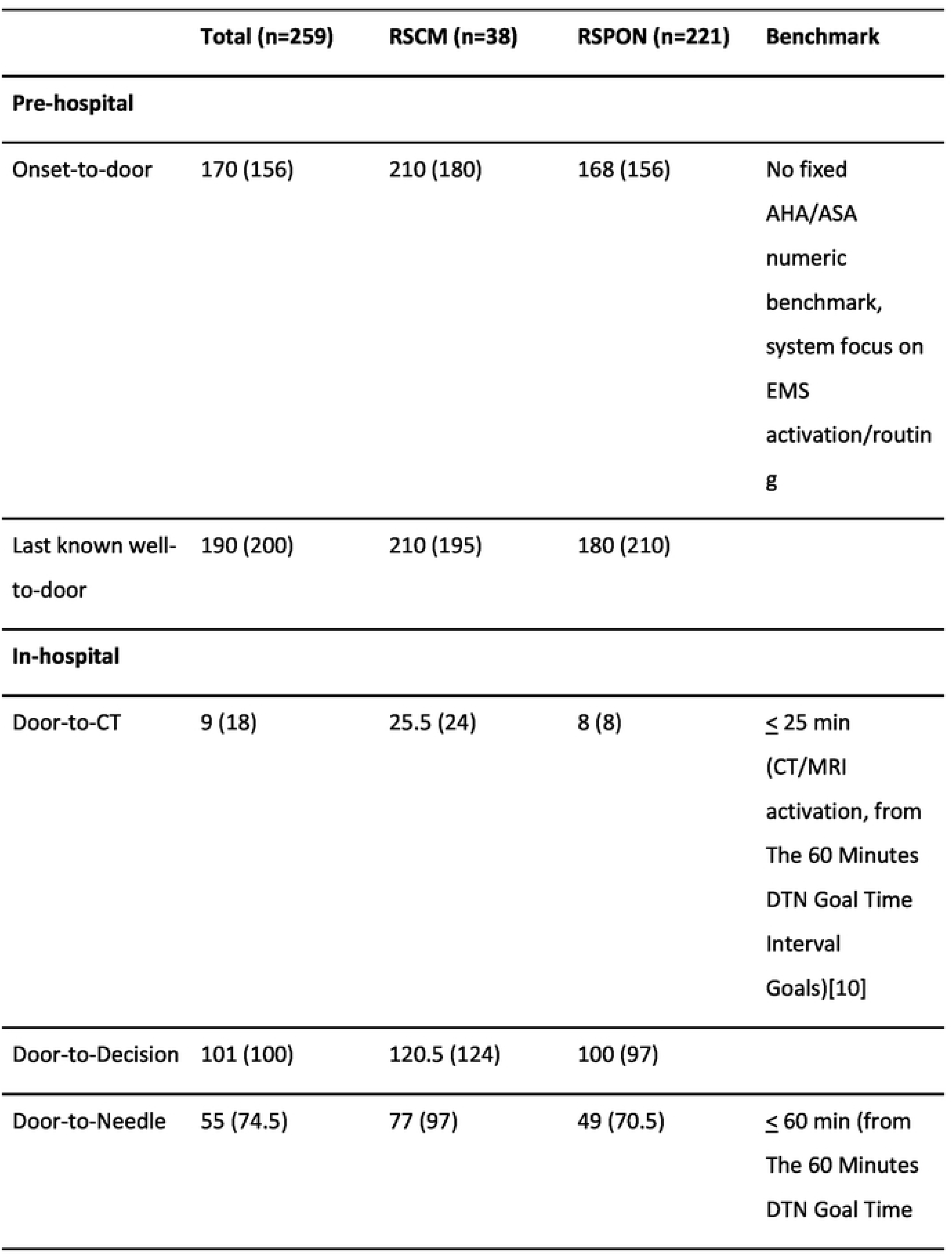
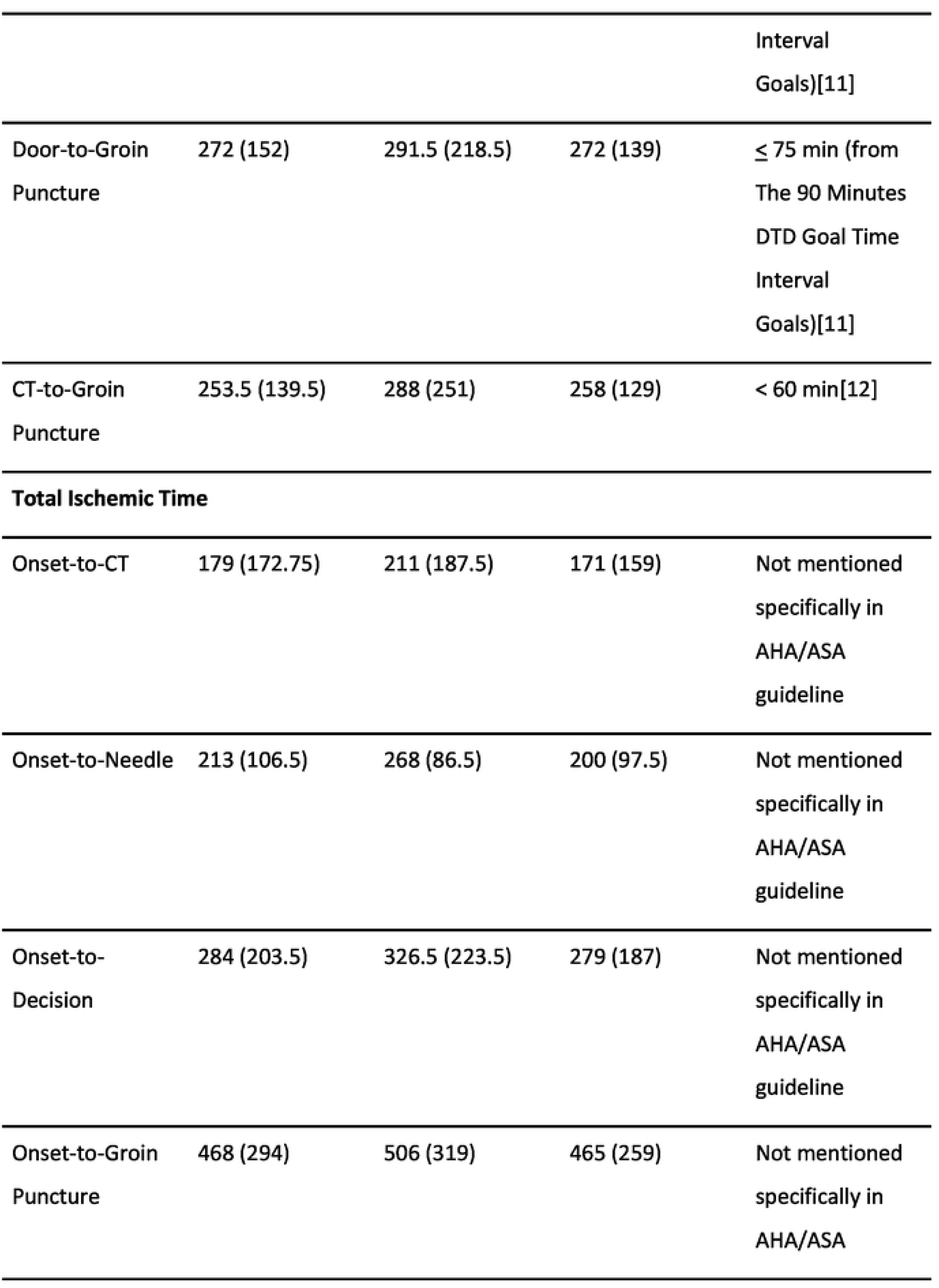
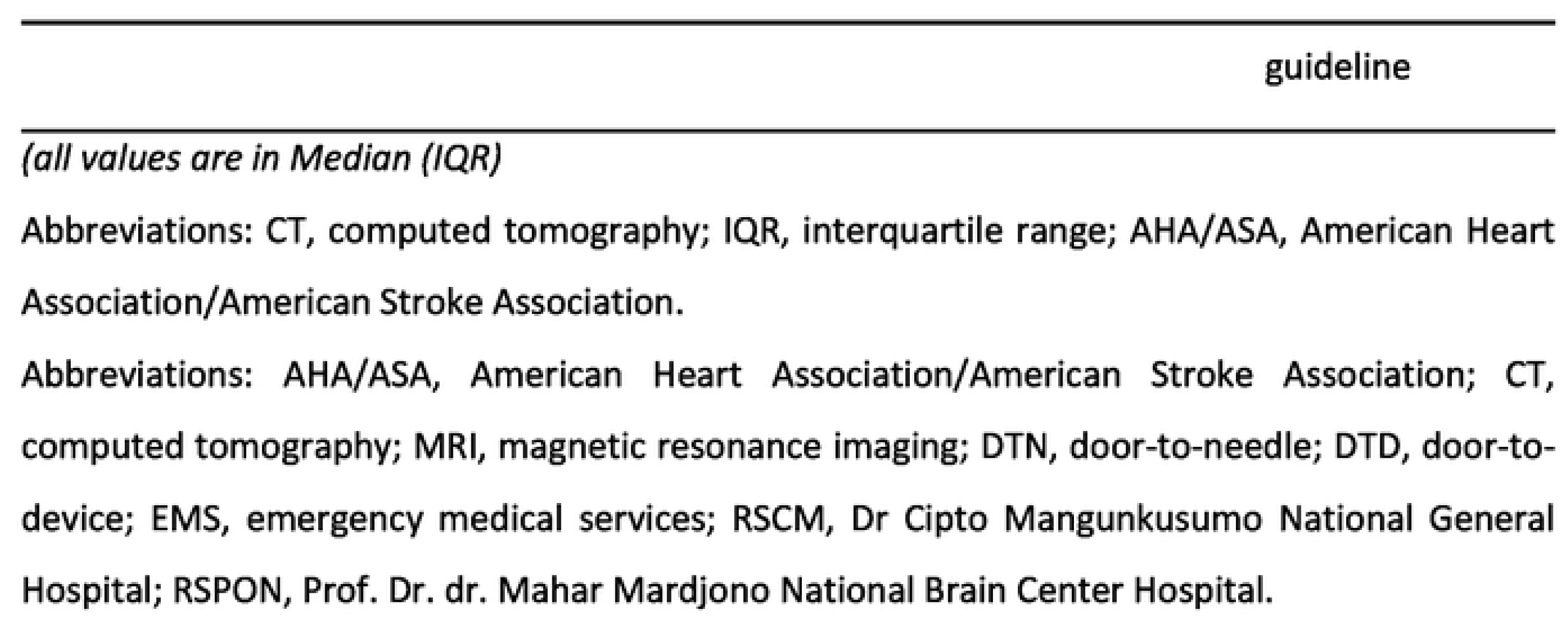
Summmary of pre-hospital and in-hospital time metrics for the total cohort and by hospital.

|  | Total (n=259) | RSCM (n=38) | RSPON (n=221) | Benchmark |
| --- | --- | --- | --- | --- |
| **Pre-hospital** |  |  |  |  |
| Onset-to-door | 170 (156) | 210 (180) | 168 (156) | No fixed<br>AHA/ASA<br>numeric<br>benchmark,<br>system focus on<br>EMS<br>activation/routin<br>g |
| Last known well-<br>to-door | 190 (200) | 210 (195) | 180 (210) |  |
| **In-hospital** |  |  |  |  |
| Door-to-CT | 9 (18) | 25.5 (24) | 8 (8) | ≤ 25 min<br>(CT/MRI<br>activation, from<br>The 60 Minutes<br>DTN Goal Time<br>Interval<br>Goals)[10] |
| Door-to-Decision | 101 (100) | 120.5 (124) | 100 (97) |  |
| Door-to-Needle | 55 (74.5) | 77 (97) | 49 (70.5) | ≤ 60 min (from<br>The 60 Minutes<br>DTN Goal Time |

|  |  |  |  | Interval<br>Goals)[11] |
| --- | --- | --- | --- | --- |
| Door-to-Groin<br>Puncture | 272 (152) | 291.5 (218.5) | 272 (139) | ≤ 75 min (from<br>The 90 Minutes<br>DTD Goal Time<br>Interval<br>Goals)[11] |
| CT-to-Groin<br>Puncture | 253.5 (139.5) | 288 (251) | 258 (129) | < 60 min[12] |
| **Total Ischemic Time** |  |  |  |  |
| Onset-to-CT | 179 (172.75) | 211 (187.5) | 171 (159) | Not mentioned<br>specifically in<br>AHA/ASA<br>guideline |
| Onset-to-Needle | 213 (106.5) | 268 (86.5) | 200 (97.5) | Not mentioned<br>specifically in<br>AHA/ASA<br>guideline |
| Onset-to-Decision | 284 (203.5) | 326.5 (223.5) | 279 (187) | Not mentioned<br>specifically in<br>AHA/ASA<br>guideline |
| Onset-to-Groin<br>Puncture | 468 (294) | 506 (319) | 465 (259) | Not mentioned<br>specifically in<br>AHA/ASA |

*(all values are in Median (IQR))*
Abbreviations: CT, computed tomography; IQR, interquartile range; AHA/ASA, American Heart Association/American Stroke Association.
Abbreviations: AHA/ASA, American Heart Association/American Stroke Association; CT, computed tomography; MRI, magnetic resonance imaging; DTN, door-to-needle; DTD, door-to-device; EMS, emergency medical services; RSCM, Dr Cipto Mangunkusumo National General Hospital; RSPON, Prof. Dr. dr. Mahar Mardjono National Brain Center Hospital.

### Institutional Evolution and Temporal Dynamics

Year-by-year analysis revealed divergent maturation patterns between the two centers. For pre-hospital coordination, the total onset-to-door trend remained relatively stable, though individual centers experienced periodic fluctuations.

In-hospital metrics demonstrated a “specialty center maturation” effect at RSPON; despite early inefficiencies in door-to-decision and door-to-groin times (peaking in 2021), the institution achieved progressive and sustained reductions in all modifiable delays from 2022 onward. Conversely, RSCM exhibited signs of system strain in the final observation year (2025), characterized by a significant sharp increase in median door-to-CT and door-to-decision times (**Figure 3**).

**Fig. 3.**
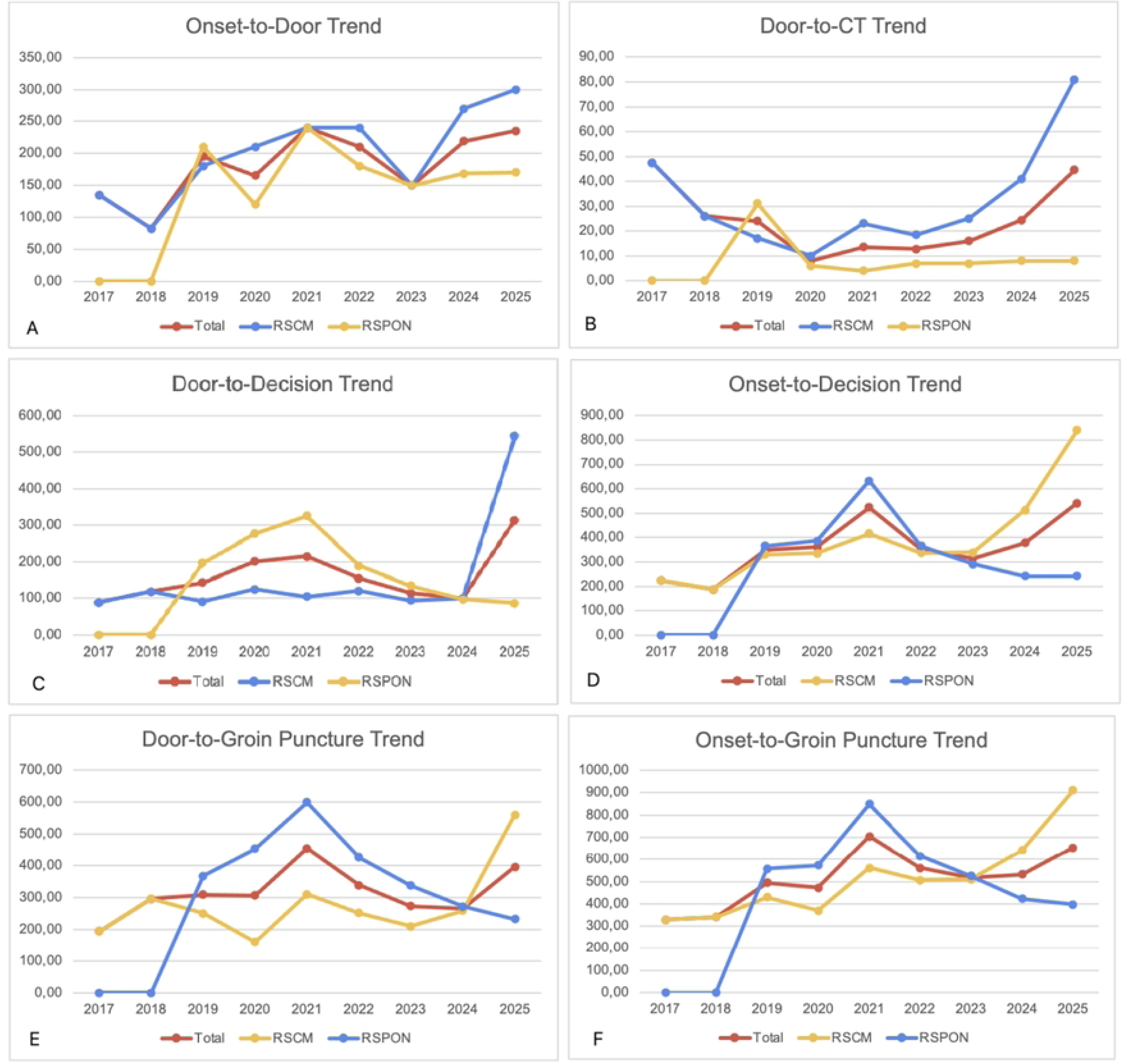
Temporal trends in key workflow intervals: (A) Onset-to-door, (B) door-to-CT, (C) door-to-Decision, (D) onset-to-decision, (E) door-to-groin, and (F) onset-to-groin.

By 2024, both centers had reached a convergence in treatment-decision efficiency, although their procedural trajectories diverged in 2025 as the dedicated neurovascular model sustained its workflow improvements despite increasing case concentration.

In **Figure 4**, RSPON demonstrates a maturation pattern with door-to-groin times progressively shortening after 2021 despite escalating MT volume. Conversely, RSCM shows relative instability in door-to-groin performance. Notably, both centers appeared to converge in 2024 (RSPON 271 vs RSCM 258 minutes) before separating sharply in 2025.

**Fig. 4.**
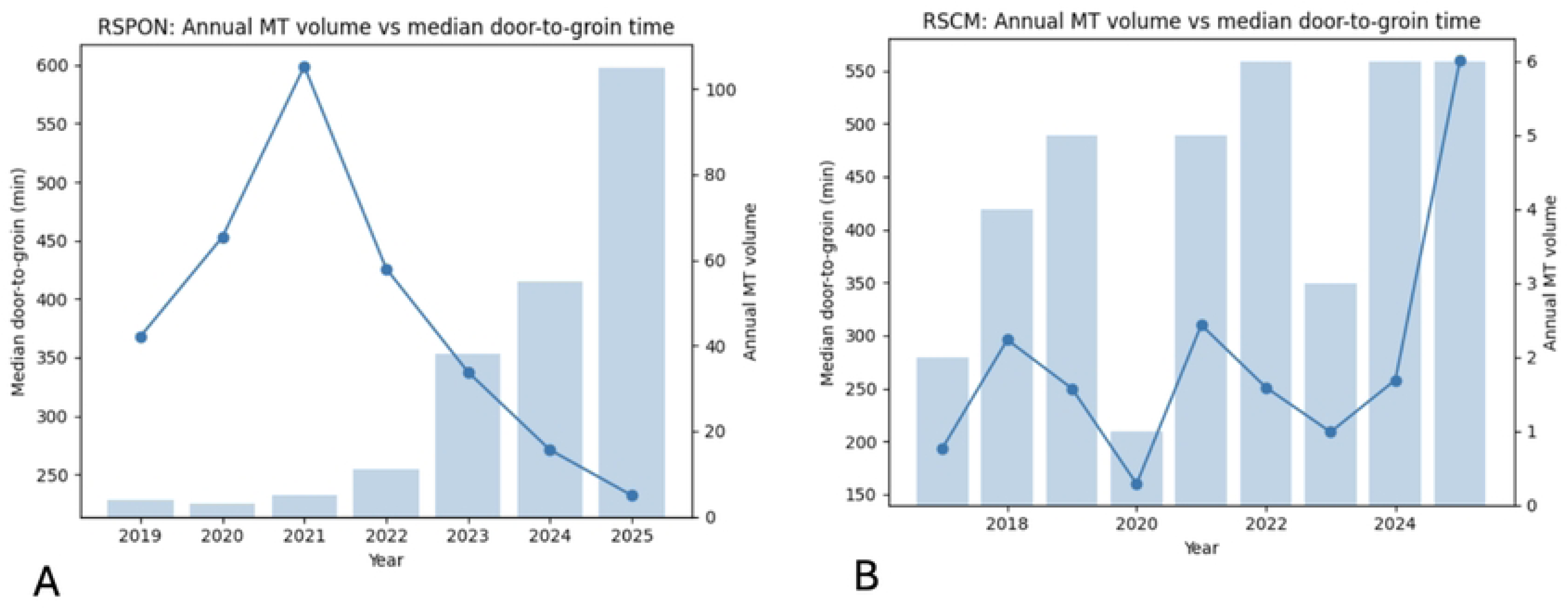
Annual MT case volume overlaid with median door-to-groin puncture time in (A) RSPON and (B) RSCM.

## Discussion

Stroke remains a major cause of death and disability in Indonesia, and national stroke services continue to evolve under the pressure of high disease burden [5]. Code stroke programs have been progressively implemented to accelerate in-hospital triage, imaging, and reperfusion workflows. At the national referral level, RSCM has achieved first intravenous thrombolysis around 2014 and endovascular therapy initiated in 2017 [7].

Our nine-year multicenter study provides a practical systems timeline of MT delivery in two tertiary centers in Jakarta. MT volumes were low and relatively stable through 2017-2022, then rose sharply in 2023-2025. This pattern is directionally consistent with disruptions during the Covid-19 era, when many regions reported reductions in stroke alerts and reperfusion activity [10–12]. Although our study cannot prove causality, the timing supports the interpretation that pandemic-related constraints and system reallocation plausibly contributed to a stagnation period before the subsequent surge.

A second macro-level signal is the prehospital ceiling. Onset-to-door times in our cohort remained broadly comparable between centers (median 168-210 minutes), and did not show the same magnitude of improvement observed in some in-hospital metrics. This mirrors broader systems literature suggesting that, while in-hospital door-to-puncture performance can improve substantially over years, prehospital time often remains relatively stagnant, reflecting persistent barriers in public symptom recognition, EMS activation, dispatcher recognition, and referral pathways [13–15]. In practical terms, even good in-hospital workflow will have a limited ceiling effect on total ischemic time if the dominant delay accumulates before arrival.

Beyond timing stability, our onset-to-door values likely reflect persistent community-level bottlenecks that are not directly modifiable by in-hospital workflow alone, including delayed symptom recognition, delayed EMS activation, traffic-related transport delays, and tiered referral pathways that prolong time-to-arrival when patients are first routed to non-EVT hospitals. These prehospital constraints are widely recognized as major determinants of total ischemic time in stroke systems-of-care, and may explain why onset-to-door intervals often improve less than in-hospital metrics over time [14].

A key contribution of this study is the comparison of two different institutional models, a dedicated neuro-focused hospital versus a general multispecialty academic hospital. RSPON showed an apparent pattern of improvement in several in-hospital workflow intervals after 2022 despite increasing MT volume. The period before 2022 is characterized by relative stagnation and greater variability in several in-hospital workflow metrics. This pattern may be consistent with workflow maturation and increasing team familiarity, although causality cannot be established in this descriptive study. As case concentration increases, teams repeat the same pathway more frequently. In addition to internal workflow maturation, the rapid increase in MT volume at RSPON may also reflect growing public and referrer awareness of RSPON as a dedicated national brain hospital, leading to preferential routing and referral of suspected cases to this center, although referral patterns were not directly measured in this study. Similar volume-efficiency associations have been reported in other systems, where higher ETV-related volumes correlate with shorter treatment times and more streamlined transfers to thrombectomy [13,16].

By contrast, RSCM showed longer median door-to-CT and door-to-decision times in 2025. However, these year-specific patterns should be interpreted cautiously because annual case volumes were smaller and medians may be unstable. A plausible explanation is institutional saturation. General emergency departments must balance multiple competing time-critical pathways such as trauma, sepsis, cardiac, and surgical emergencies, and without highly standardized neurovascular escalation, stroke alert may become vulnerable to congestion and resource competition. This interpretation aligns with broader registry evidence that more advanced stroke center structures tend to achieve key performance targets more reliably than less specialized settings. This reflects differences in infrastructure, staffing models, and workflow governance [17].

These between-center differences were also reflected in onset-to-groin times. RSPON achieved shorter onset-to-groin puncture times than RSCM. Since earlier reperfusion is strongly associated with better outcomes and treatment effect, the observed between-center differences may be clinically relevant [18,19], but this study was not designed to evaluate outcome effects.

Across the pooled cohort, early imaging was relatively rapid, particularly at RSPON (median door-to-CT 8 minutes). Door-to-CT is often used as a front-end marker of ED triage-to-imaging throughput. International stroke quality measurement programs commonly benchmark brain imaging within 25 minutes of arrival [20]. In our dataset, this front-end step appears achievable, but also sensitive to congestion, which is consistent with the late rise in door-to-CT at RSCM.

Door-to-decision was prolonged in both centers, indicating a major decision gap. Conceptually, the interval blends cognitive work such as imaging interpretation, eligibility information, and weighing risk/benefit with administrative coordination such as mobilizing multidisciplinary consensus, family consent, and financial logistics. Contemporary guidelines emphasize that thrombectomy evaluation and treatment should occur as rapidly as possible, therefore, systematically tracking the intermediate metrics beyond door-to-CT has become important for governance and quality improvement [21]. The implication is that hospital should treat decision as an operational output of a pathway, not merely an individual clinician’s step. This “decision gap” is frequently amplified by several units where communication delays can accumulate. A practical approach described in other stroke quality-improvement programs is to formalize a stroke coordinator role to drive parallel processing. A study from Taiwan included stroke nursing specialists and used trained study nurses/stroke case managers as part of the implementation, with significant improvements in key acute stroke measures [22].

The most striking micro-level findings is that the dominant in-hospital time burden occurs after imaging. CT-to-groin puncture medians remained very long (258-288 minutes), driving door-to-groin medians (272-292 minutes). This procedural lag typically reflects cathlab/angiosuite availability, team mobilization, anesthesia readiness, patient transport between units, and the efficiency of CT-to-angio transitions [23]. Multiple quality-improvement studies and pathway redesign trials have shown that this segment is highly modifiable using strategies such as parallel processing, pre-notification of the EVT team, direct-to-angiosuite models for selected patients, and streamlined transport/handoff protocols [24,25].

Clinically, the importance of shortening post-imaging delays is supported by patient-level evidence showing that longer onset-to-puncture times reduce the probability of favorable outcomes and may reduce the likelihood and quality of reperfusion [19,26]. In other words, even when door-to-CT is excellent, prolonged CT-to-groin can erase the benefit of early recognition and fast imaging.

Taken together, despite the different institutional profiles of two hospitals, both pathways demonstrate clear room for improvement at the micro level. The consistent dominance of post-imaging delays across centers suggests that focusing on decision and endovascular readiness processes may yield the greatest gains in treatment timelines.

At the macro level, improving total ischemic time will require strengthening pre-hospital system, such as symptom recognition campaigns, EMS dispatcher stroke triage, and routing policies that reduce interfacility transfer delays [14,15,27]. At the meso level, the divergence between a neuro-focused pathway and a general-hospital pathway suggested that governance, resource, and standardized escalation may matter as much as experience.

Registry data indicating better target achievement in more advanced stroke center models supposrts investments in thrombectomy-capable infrastructures [7]. At the micro level, the most actionable bottlenecks in our cohort sit in door-to-decision and especially CT-to-groin. Interventions should prioritize post-imaging parallelization (consent, anesthesia planning, angiosuite activation) and define time-to-ready milestones that can be audited continuously [25,28].

This study has several limitations. This retrospective study relies on medical-record timestamps, so workflow metrics may be affected by incomplete or inconsistent documentation and variation in operational definitions across centers and years. Annual trend estimates should be interpreted cautiously because small year-by-year case numbers particularly in the lower volume center can make medians sensitive to outliers. In addition, we did not account for temporal changes in referral pathways, transfer status, case mix, or imaging-selection practices, all of which may influence workflow intervals independently of process efficiency. The analysis includes only two tertiary hospitals in a single urban setting, limiting generalizability to other Indonesian regions. Finally, because this descriptive study focused on time metrics, it was not designed to assess clinical outcomes or to quantify the outcome impact of specific workflow intervals within this cohort.

## Conclusion

In summary, this study highlights several priority areas for strengthening stroke systems in Indonesia. The persistently prolonged post-imaging intervals, particularly the CT-to-groin segment, which remained the dominant in-hospital delay, underscore the need for hospital-level policies that streamline decision-making, anesthesia readiness, and inter-unit transport. The relative stagnation of onset-to-door times further indicates that national reforms must extend beyond hospitals to include EMS dispatcher stroke triage and public education to reduce recognition and activation delays. Together, these findings support a coordinated policy approach that integrates prehospital strengthening, hospital workflow redesign, and strategic investment in thrombectomy-capable infrastructures. Future research should link workflow intervals to clinical outcomes to quantify the functional impact of delays in the Indonesian context, examine how organizational structures, staffing models, and governance mechanisms influence performance, and establish a national thrombectomy registry to enable continuous monitoring, benchmarking, and quality-improvement across diverse hospital settings.

## Data Availability

The de-identified minimal dataset underlying the findings of this study is publicly available at https://docs.google.com/spreadsheets/d/1TyYJlO8QIIHONsgCTn7K-GwYzzQzM9vTyi_Mq4-Q3PY/edit?gid=1583759278#gid=1583759278

## Statements

## Acknowledgement

None

## Statement of Ethics

### Study approval statement

This retrospective secondary analysis was conducted under an approved parent research protocol. Ethical approval was obtained from the Ethics Committee of the Faculty of Medicine, Universitas Indonesia–Cipto Mangunkusumo Hospital (No. KET- 402/UN2.F1/ETIK/PPM.00.02/2026; Protocol No. 26-05-0804; approved on June 12, 2026); and the Health Research Ethics Committee of Prof. Dr. dr. Mahar Mardjono National Brain Center Hospital (No. DP.04.03/D.XXIII.9/56/2025, with protocol amendment approval No. DP.04.03/D.XXIII.9/93/2025). The requirement for individual informed consent was waived because this study involved the retrospective review of existing medical records. All patient data were de-identified before analysis.

## Author Contributions

All authors contribute equally in this research.

## Data Availability Statement

The data that support the findings of this study are not publicly available due to privacy reasons but are available from the corresponding author upon reasonable request.

## References

[1] Turc G, Bhogal P, Fischer U, Khatri P, Lobotesis K, Mazighi M, et al. European Stroke Organisation (ESO) - European Society for Minimally Invasive Neurological Therapy (ESMINT) Guidelines on Mechanical Thrombectomy in Acute Ischemic Stroke. J Neurointerv Surg 2023;15. 10.1136/neurintsurg-2018-014569.

[2] Legere B, Mohamed A, Elsherif S, Saqqur R, Schoenfeld D, Slebonick AM, et al. Success with incrementally faster times to endovascular therapy (SWIFT-EVT): A systematic review and meta-analysis. Journal of Stroke and Cerebrovascular Diseases 2024;33. 10.1016/j.jstrokecerebrovasdis.2024.107964.

[3] Saver JL, Goyal M, Van Der Lugt A, Menon BK, Majoie CBLM, Dippel DW, et al. Time to Treatment With Endovascular Thrombectomy and Outcomes From Ischemic Stroke: A Meta-analysis. JAMA 2016;316:1279–89. 10.1001/jama.2016.13647.

[4] Ruff IM, Ali SF, Goldstein JN, Lev M, Copen WA, McIntyre J, et al. Improving Door-to-Needle Time: A Single Center Validation of the Target Stroke Hypothesis. Stroke 2014;44:504–8. 10.1161/STROKEAHA.

[5] Venketasubramanian N, Yudiarto FL, Tugasworo D. Stroke Burden and Stroke Services in Indonesia. Cerebrovasc Dis Extra 2022;12:53–7. 10.1159/000524161.

[6] Rasyid A, Kurniawan M, Mesiano T, Hidayat R, Rilianto B, Harris S. Performance of door-to-CT time of code stroke in Indonesian tertiary referral center hospital. Egyptian Journal of Neurology, Psychiatry and Neurosurgery 2022;58. 10.1186/s41983-022-00583-6.

[7] Mesiano T, Kurniawan M, Saputri KM, Hidayat R, Permana AP, Rasyid A, et al. Endovascular Treatment in Acute Ischemic Stroke Adoption and Practice: A Single-Center Indonesian Experience. Cerebrovasc Dis Extra 2021;11:72–6. 10.1159/000517183.

[8] Kurniawan M, Mulya Saputri K, Mesiano T, Yunus RE, Permana AP, Sulistio S, et al. Efficacy of endovascular therapy for stroke in developing country: A single-centre retrospective observational study in Indonesia from 2017 to 2021. Heliyon 2024;10. 10.1016/j.heliyon.2023.e23228.

[9] Ameen D, Dewey HM, Khalil H. Strategies to reduce delays in delivering mechanical thrombectomy for acute ischaemic stroke – an umbrella review. Frontiers in Neurology 2024;15. 10.3389/fneur.2024.1390482.

[10] July J, Pranata R. Impact of the Coronavirus Disease Pandemic on the Number of Strokes and Mechanical Thrombectomies: A Systematic Review and Meta-Analysis. Journal of Stroke and Cerebrovascular Diseases 2020;29:105185. 10.1016/j.jstrokecerebrovasdis.2020.105185.

[11] Nogueira RG, Abdalkader M, Qureshi MM, Frankel MR, Mansour OY, Yamagami H, et al. Global impact of COVID-19 on stroke care. International Journal of Stroke 2021;16:573–84. 10.1177/1747493021991652.

[12] Nguyen TN, Qureshi MM, Klein P, Yamagami H, Mikulik R, Czlonkowska A, et al. Global Impact of the COVID-19 Pandemic on Stroke Volumes and Cerebrovascular Events. Neurology 2023;100:E408–21. 10.1212/WNL.0000000000201426.

[13] Sun C, Zaidat OO, Castonguay AC, Veznedaroglu E, Budzik RF, English J, et al. A Decade of Improvement in Door-to-Puncture Times for Mechanical Thrombectomy But Ongoing Stagnation in Prehospital Care. Stroke: Vascular and Interventional Neurology 2022;3:e000561. 10.1161/svin.122.000561.

[14] Soto-Camara R, Gonzales-Bernal J, Aguilar-Parra JM, Trigueros R, Lopez-Liria R, Gonzales- Santos J. Factors related to prehospital time in caring for patients with stroke. Emergencias 2021;33:454–63.

[15] Zachrison KS, Nielsen VM, De La Ossa NP, Madsen TE, Cash RE, Crowe RP, et al. Prehospital Stroke Care Part 1: Emergency Medical Services and the Stroke Systems of Care. Stroke 2023;54:1138–47. 10.1161/STROKEAHA.122.039586;WGROUP:STRING:PUBLICATION.

[16] van Meenen LCC, den Hartog SJ, Groot AE, Emmer BJ, Smeekes MD, Siegers A, et al. Relationship between primary stroke center volume and time to endovascular thrombectomy in acute ischemic stroke. Eur J Neurol 2021;28:4031. 10.1111/ene.15107.

[17] Raychev R, Sun JL, Schwamm L, Smith EE, Fonarow GC, Messé SR, et al. Performance of Thrombectomy-Capable, Comprehensive, and Primary Stroke Centers in Reperfusion Therapies for Acute Ischemic Stroke: Report From the Get With The Guidelines-Stroke Registry. Circulation 2023;148:2019–28. 10.1161/CIRCULATIONAHA.123.066114.

[18] Saver JL, Goyal M, Van Der Lugt A, Menon BK, Charles;, Majoie BLM, et al. Time to Treatment With Endovascular Thrombectomy and Outcomes From Ischemic Stroke: A Meta-analysis Supplemental content. JAMA 2016;316:1279–88. 10.1001/jama.2016.13647.

[19] Bourcier R, Goyal M, Liebeskind DS, Muir KW, Desal H, Siddiqui AH, et al. Association of Time From Stroke Onset to Groin Puncture With Quality of Reperfusion After Mechanical Thrombectomy: A Meta-analysis of Individual Patient Data From 7 Randomized Clinical Trials. JAMA Neurol 2019;76:405. 10.1001/jamaneurol.2018.4510.

[20] Kelly AG, Hellkamp AS, Olson D, Smith EE, Schwamm LH. Predictors of rapid brain imaging in acute stroke: Analysis of the get with the guidelines-stroke program. Stroke 2012;43:1279–84. 10.1161/STROKEAHA.111.626374.

[21] Prabhakaran S, Gonzalez NR, Zachrison KS, Adeoye O, Alexandrov AW, Ansari SA, et al. 2026 Guideline for the Early Management of Patients With Acute Ischemic Stroke: A Guideline From the American Heart Association/American Stroke Association. Stroke 2026;57:0–00. 10.1161/STR.0000000000000513.

[22] Hsieh FI, Jeng JS, Chern CM, Lee TH, Tang SC, Tsai LK, et al. Quality improvement in acute ischemic stroke care in Taiwan: The breakthrough collaborative in stroke. PLoS One 2016;11. 10.1371/journal.pone.0160426.

[23] Schregel K, Behme D, Tsogkas I, Knauth M, Maier I, Karch A, et al. Effects of Workflow Optimization in Endovascularly Treated Stroke Patients – A Pre-Post Effectiveness Study. PLoS One 2016;11:e0169192. 10.1371/JOURNAL.PONE.0169192.

[24] Settecase F, McCoy DB, Darflinger R, Alexander MD, Cooke DL, Dowd CF, et al. Improving mechanical thrombectomy time metrics in the angiography suite: Stroke cart, parallel workflows, and conscious sedation. Interventional Neuroradiology 2018;24:168–77. 10.1177/1591019917742326.

[25] Pfaff JAR, Schönenberger S, Herweh C, Ulfert C, Nagel S, Ringleb PA, et al. Direct Transfer to Angio-Suite Versus Computed Tomography-Transit in Patients Receiving Mechanical Thrombectomy: A Randomized Trial. Stroke 2020;51:2630–8. 10.1161/STROKEAHA.120.029905.

[26] Saver JL, Goyal M, Van Der Lugt A, Menon BK, Majoie CBLM, Dippel DW, et al. Time to Treatment With Endovascular Thrombectomy and Outcomes From Ischemic Stroke: A Meta-analysis. JAMA 2016;316:1279–88. 10.1001/JAMA.2016.13647.

[27] Riegler C, Behrens JR, Gorski C, Angermaier A, Kinze S, Ganeshan R, et al. Time-to-care metrics in patients with interhospital transfer for mechanical thrombectomy in north-east Germany: Primary telestroke centers in rural areas vs. primary stroke centers in a metropolitan area. Front Neurol 2023;13:1046564. 10.3389/fneur.2022.1046564.

[28] Rangel I, Palmisciano P, Vanderhye VK, El Ahmadieh TY, Wahood W, Demaerschalk BM, et al. Optimizing Door-to-Groin Puncture Time: The Mayo Clinic Experience. Mayo Clin Proc Innov Qual Outcomes 2022;6:327–36. 10.1016/j.mayocpiqo.2022.05.009.

